# An R83W mutation in Rab3A causes autosomal-dominant cerebellar ataxia

**DOI:** 10.1101/2025.07.16.25330541

**Authors:** Ryosuke Miyamoto, Ayuko Sakane, Hiroyuki Morino, Kodai Kume, Tomoyasu Matsubara, Tatsuya Fukumoto, Yusuke Osaki, Ryosuke Oki, Kenta Hanada, Konoka Tachibana, Masahito Nakataki, Yoshihiko Nishida, Yuji Takahashi, Kenji Mizuguchi, Shigeo Murayama, Yuko Saito, Hideshi Kawakami, Yoshimi Takai, Takuya Sasaki, Yuishin Izumi

## Abstract

Spinocerebellar ataxias (SCAs) are a group of progressive neurodegenerative disorders caused by pathogenic variants in more than 40 genes with diverse cellular functions. In this study, we identified the c.247C>T p.(Arg83Trp) variant in *RAB3A*, encoding a small GTPase involved in membrane-associated regulated exocytosis, in two families with cerebellar ataxia. Affected individuals presented with adult-onset, gradually progressive cerebellar symptoms, often accompanied by mild gait spasticity and tremors. Variable features of neurodevelopmental disorders were also observed. Brain MRI consistently revealed cerebellar atrophy, often accentuated in the vermis, and neuropathological examinations demonstrated diffuse cerebellar cortical degeneration. Functionally, the R83W mutation lies within the conserved switch II region of Rab3A, a domain critical for effector interaction. Although the mutant Rab3A R83W retained GTP-binding affinity, it failed to bind the key effector proteins RIM1 and Rabphilin-3A, highlighting the functional importance of R83 in effector complex formation, as supported by structural analysis. In PC12 cells, the R83W mutant exhibited diffuse cytoplasmic localization, in contrast to the vesicle- and neurite-tip localization of the wild-type and GTP-bound Rab3A mutant. The concordant localization pattern of R83W and GDP-bound Rab3A mutants suggests that R83W-induced mislocalization results from a failure to engage downstream effector proteins. In frozen sections of the mouse cerebellum, Rab3A was predominantly localized to parallel fiber terminals and was absent from postsynaptic Purkinje cells. These findings suggest that disruption of the interaction between Rab3A and its effector proteins may underlie disease pathogenesis, possibly involving presynaptic dysfunction at parallel fiber–Purkinje cell synapses mediated by the Rab3A–RIM1 complex.

## Introduction

Spinocerebellar ataxias (SCAs) are a genetically diverse group of neurodegenerative disorders predominantly affecting the cerebellum, leading to progressive motor incoordination and other neurological deficits, such as peripheral neuropathy, tremors, pyramidal tract signs, and cognitive impairment [1]. To date, over 40 genetically distinct SCA subtypes have been identified, and the increasing implementation of whole-exome sequencing has steadily driven the identification of nonrepeat mutations, such as point mutations in SCA42 (MIM: 616795) [2, 3] and short insertions/deletions in SCA48 (MIM: 618093) [4]. The pathophysiology triggered by mutated SCA genes typically involves several common mechanisms, including polyglutamine toxicity, proteotoxicity, RNA toxicity, channel dysfunction, impaired bioenergetics, and loss of nuclear integrity [1]; however, accumulating evidence highlights that synaptic dysfunction is a shared underlying pathological mechanism in various SCAs, and that understanding the molecular basis of synaptic function and its disruption is important for identifying potential therapeutic targets [5].

Intracellular membrane trafficking is a fundamental process in neuronal function and is essential for neurotransmitter release and synaptic plasticity [6]. This process is tightly regulated by a large family of small GTPases known as Rab proteins [7]. The Rab3 subfamily, consisting of four closely related members—Rab3A, Rab3B, Rab3C, and Rab3D—is particularly associated with secretory vesicles and granules and plays a crucial role in regulated exocytosis. These proteins are expressed at the highest levels in the brain and endocrine tissues and appear to be localized on exocytic vesicles [6, 7].

Rab3A is the most abundant Rab isoform in the brain, where it is primarily localized on synaptic vesicles (SVs) [8]. It functions as a molecular switch, cycling between an inactive GDP-bound state in the cytosol and an active GTP-bound state associated with vesicle membranes. This cyclical activation, coupled with membrane association, is thought to enable spatial and temporal control of Rab3A activity in vesicle transport. This crucial function as a molecular switch, particularly in the active GTP-bound state, is mediated through interactions with specific downstream effector proteins, particularly RIM1 [9] and Rabphilin-3A [10], which are known to regulate distinct steps of vesicular transport and release, including docking and priming at the presynaptic active zone. Consequently, proper functioning and interaction of Rab3A with binding partners critically contribute to cellular homeostasis, particularly in the nervous system [6]. Disruptions in Rab3A function or genetic defects in its effectors and regulators, such as Rab3 GTPase-activating protein and Rab GDP dissociation inhibitor, can lead to impaired cellular processes, with some contributing to the pathogenesis of neurodevelopmental and neurodegenerative diseases [11–14].

In this study, we identified a pathogenic *RAB3A* variant, c.247C>T p.(Arg83Trp), in two families with adult-onset cerebellar ataxia and variable features of neurodevelopmental disorders. Notably, the R83W mutation disrupted the association of Rab3A with the effector proteins RIM1 and Rabphilin-3A while preserving the GTP-binding affinity. The R83W mutant exhibited a diffuse cytoplasmic localization, suggesting that impaired effector protein binding leads to mislocalization of Rab3A and potentially disrupts presynaptic neurotransmission. Furthermore, in frozen sections of the mouse cerebellum, Rab3A was predominantly localized to parallel fiber (PF) terminals and was absent from Purkinje cells (PCs). These findings suggest that disruption of the interaction between Rab3A and its effector proteins may contribute to disease pathogenesis, possibly involving presynaptic dysfunction at PF terminals mediated by the Rab3A–RIM1 complex. Elucidation of this specific molecular pathway provides fundamental insights into the mechanisms underlying synaptic dysfunction in this newly identified form of SCA and potentially other neurodevelopmental disorders where synaptic impairment is a common feature.

## Results

### Clinical phenotype

The clinical characteristics of the affected individuals from Families T and K (Fig. 1A) are summarized in Table 1. The mean age of onset was 39.0 years. All 9 affected individuals presented a gait instability as the initial symptom, followed by a gradually progressive disease course. The cardinal clinical features included truncal- predominant ataxia, wide-based gait, slurred speech, and horizontal gaze nystagmus. Among the extracerebellar signs, mild gait spasticity with lower limb hyperreflexia was common. Tremors involving the neck, trunk, and upper limbs were observed in 5 individuals. In individual T-II:2, neck tremors were present at a frequency of 2–3 Hz and were exacerbated during stance or gait. An extensor plantar response was observed in 2 individuals. Moreover, variable neurodevelopmental disorders/features, ranging from autism spectrum disorder (ASD) traits to intellectual disabilities, were noted in 5 individuals. No affected individuals exhibited sensory, urinary, or bowel dysfunction.

**Figure 1.**
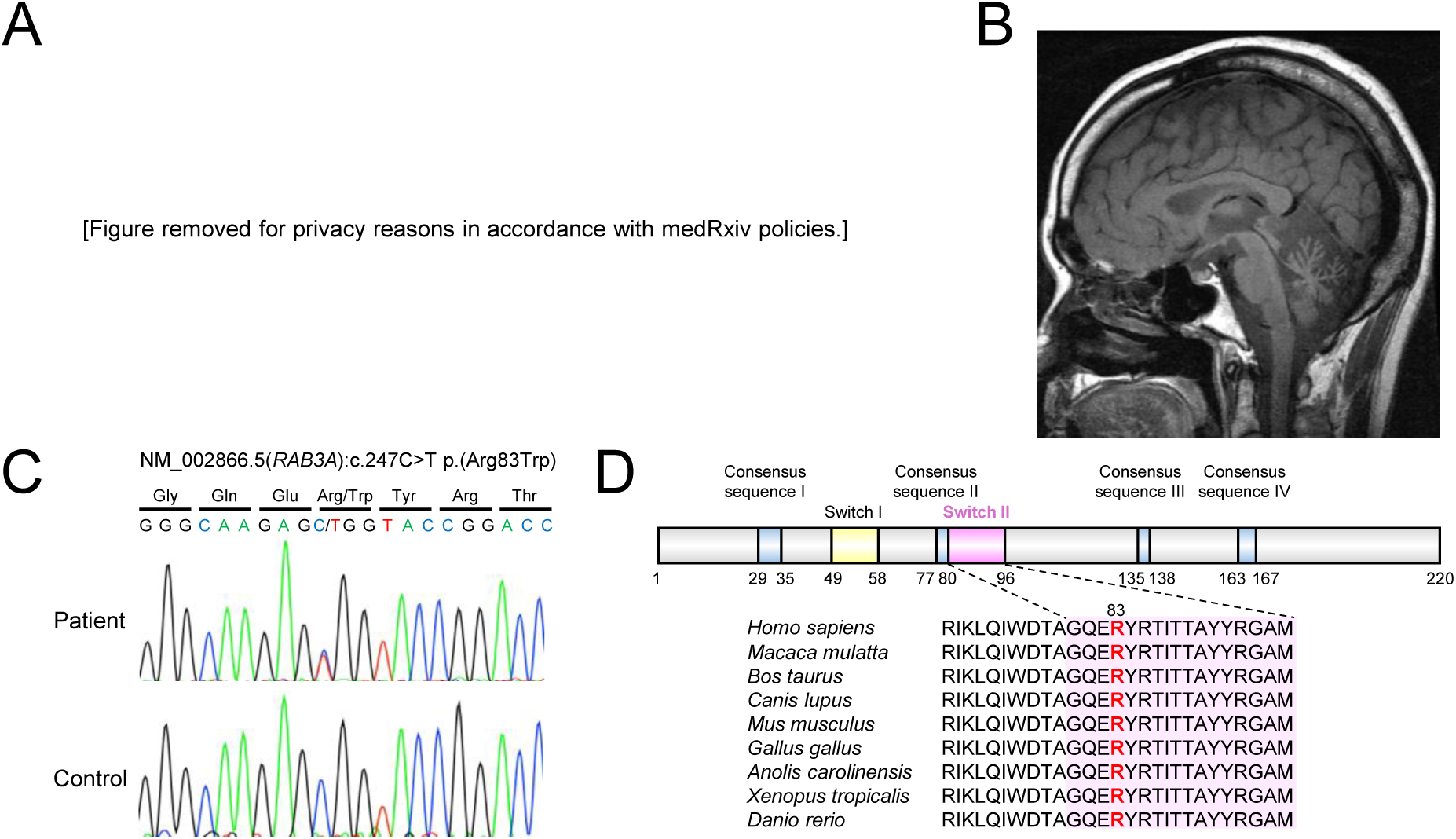
Identification of a mutation in *RAB3A* as a cause of SCA. (A) Pedigree structures of Families T and K. Asterisks indicate individuals who underwent whole- exome sequencing and linkage analysis. Not all family members are shown in the pedigrees. Full data are available from the corresponding author upon reasonable request. (B) T1-weighted MR image of the index patient (T-II:2) demonstrating cerebellar vermian atrophy. (C) Sanger sequencing chromatogram of the identified mutation. (D) Structural and evolutionary characterization of Rab3A. The R83 residue is located within the switch II region and is adjacent to consensus sequence II. Both regions are highly conserved across species.

**Table 1.** Clinical characteristics of the affected individuals.

|  | A-I:2 | A-II:1 | A-II:2 | A-II:5 | A-II:7 | A-III:1 | A-III:3 | A-III:4 | B-II:2 |
| --- | --- | --- | --- | --- | --- | --- | --- | --- | --- |
| Sex | Female | Female | Female | Female | Male | Male | Male | Male | Female |
| Symptom at onset | Unstable gait | Unstable gait | Unstable gait | Unstable gait | Unstable gait | Unstable gait | Unstable gait | Unstable gait | Unstable gait |
| SARA | N/A | 33 | 23 | 17 | 11 | 14.5 | 19 | 10.5 | 25 |
| Ambulatory status | Unable to walk | Unable to walk | Walker | Cane | Independent | Independent | Cane | Independent | Wheelchair |
| Ataxia | + | Trunk > Limb | Trunk > Limb | Trunk > Limb | Trunk > Limb | Trunk > Limb | Trunk > Limb | Trunk > Limb | Trunk > Limb |
| Gait | N/A | Wide-based | Wide-based, spastic | Wide-based, spastic | Wide-based, spastic | Wide-based | Wide-based, spastic | Wide-based, spastic | Wide-based |
| Speech | N/A | Slurred | Slurred | Slurred | Slurred | Slurred | Slurred, explosive | Slurred | Slurred, explosive |
| Ocular movement | N/A | SP, HGN | SP, HGN | SP, HGN | SP, HGN | HGN | SP, SID, HGN | SP, HGN | SP, HGN |
| Tremor | +(Neck) | +(Neck) | +(Neck, trunk) | – | +(UL) | – | – | – | +(Neck, trunk) |
| Reflexes (UL/LL) | N/A | Normal/Normal | Normal/Hyper | Hyper/Hyper | Hyper/Hyper | Normal/Normal | Normal/Hyper | Normal/Hyper | Hypo/Hypo |
| Extensor plantar response | N/A | – | + | – | + | – | – | – | – |
| Sensory impairments | N/A | – | – | – | – | – | – | – | – |
| Autonomic impairments | N/A | – | – | – | – | – | – | – | – |
| Neurodevelopmental disorders/features | N/A | N/A | N/A | +(ASD traits) | +(Mild ID) | +(BIF) | +(ASD traits) | +(ASD) | – |
| MMSE | N/A | 22 | 22 | 28 | 15 | 24 | 30 | 30 | N/A |
| Cerebellar atrophy | N/A | + | + | + | + | + | + | + | + |
| Other findings |  |  | Normal NCS |  | Rt. temporal atrophy | FIQ 72 (WAIS-III) |  |  |  |

N/A: not assessed; SARA: Scale for the Assessment and Rating of Ataxia; SP: saccadic pursuit; HGN: horizontal gaze nystagmus; SID: saccade
initiation delay; UL: upper limb; LL: lower limb; Hyper: hyperactive; Hypo: hypoactive; ASD: autism spectrum disorder; ID: intellectual disability;
BIF: borderline intellectual functioning; MMSE: Mini-Mental State Examination; NCS: nerve conduction study; Rt: right; FIQ: full-scale intelligence
quotient; WAIS-III: Wechsler Adult Intelligence Scale - Third Edition. Full data are available from the corresponding author upon reasonable request.

Brain MRI of all the evaluated individuals revealed cerebellar atrophy, often accentuated in the vermis (Fig. 1B, Fig. S1). Routine blood tests revealed no significant abnormalities.

### Genetic analysis

Genome-wide linkage analysis in Family T did not identify a unique associated locus due to the limitations of the small sample size and pedigree structure but did display multiple peaks (Fig. S2). The three largest regions among those with the highest LOD score (1.23) were found on chromosomes 7, 9, and 19. Next, whole-exome sequencing was performed on 5 affected individuals and 1 unaffected individual.

Variants in the heterozygous state were filtered according to their functional class (nonsynonymous, frameshift, stop, or splicing), frequency in publicly available genomic databases and in-house exome databases, and predicted pathogenicity by CADD (phred > 20). These criteria reduced the number of candidate variants to 44 in the proband and only one missense variant in *RAB3A*, NM_002866.5:c.247C>T p.(Arg83Trp), was segregated with the ataxia phenotype (Fig. 1C, Table S1). Segregation was subsequently confirmed via Sanger sequencing in 11 individuals, including 7 affected and 4 unaffected individuals. The *RAB3A* gene is located at 19p13.11 and was one of the candidate regions suggested by linkage analysis. The R83W variant was predicted to be deleterious by CADD (phred = 26.5), SIFT, Polyphen-2, and MutationTaster. The altered amino acid is highly conserved across species and is located in the conserved switch II region, which is sensitive to the GDP/GTP-binding state of Rab3A [15] (Fig. 1D). Furthermore, we screened 320 patients with spinocerebellar ataxia for *RAB3A* variants using amplicon sequencing and identified the same c.247C>T p.(Arg83Trp) variant in one sporadic case (patient K-II:2).

### Neuropathological findings

An autopsy was performed for patient T-II:1. The brain weighed 1,029 g before fixation. Macroscopic examination revealed a markedly atrophic cerebellum (Fig. 2A), and this change was predominant in the vermis and the superior parts of the hemispheres (Fig. 2B). Microscopically, the cerebellum exhibited diffuse cortical degeneration, including thinning of the molecular layers, severe loss of PCs accompanied by Bergmann gliosis, loss of the granular cell layers, and gliosis in the white matter (Fig. 2C). Many empty basket fibers were observed in the PC layers. In addition, a few cactus-like swollen neurites in the molecular layers and a few torpedoes in the granular cell layers were also observed (Fig. 2D). The dentate nuclei showed gliosis, and the surrounding white matter exhibited myelin pallor, although the dentate neurons were relatively preserved. The inferior olivary nuclei presented mild neuronal loss, predominantly in the dorsal part (Fig. 2E and 2F). Other areas of the brain, the spinal cord, and the dorsal root ganglia exhibited no apparent abnormal changes.

**Figure 2.**
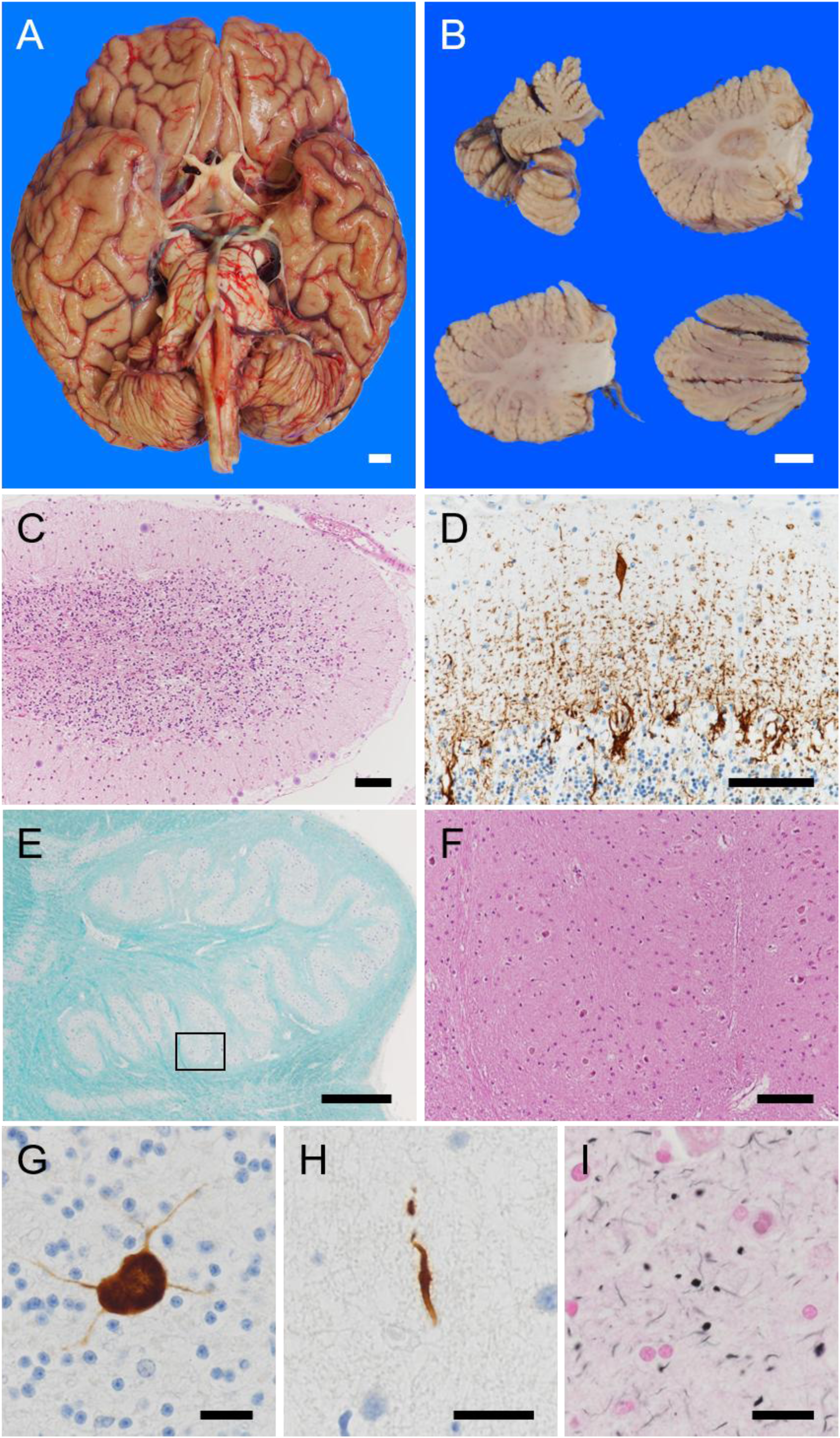
Neuropathological findings. Macroscopically, the cerebellum was markedly atrophic (A), and sagittal sections of the left cerebellum revealed atrophy of folia predominantly in the vermis and the superior parts of the hemispheres (B). The cerebellar cortex of the vermis showed severe neuronal loss with gliosis throughout all layers (C). Immunostaining for phosphorylated neurofilaments revealed many empty basket fibers and a cactus-like swollen neurite (D). Mild neuronal loss with gliosis was observed in the dorsomedial part of the inferior olive nucleus (E: low-power field with Klüver–Barrera staining; F: high-power field shown in the square in E with hematoxylin and eosin staining). A medium-sized multipolar neuron in the granular layer of the vermis exhibited a p62-immunoreactive intracytoplasmic inclusion (G). Scattered phosphorylated TDP43-immunoreactive threads were observed in the molecular layers of the cerebellum (H). Gallyas–Braak silver impregnation staining of the amygdala revealed many argyrophilic grains (I). Scale bars: 1 cm (A, B), 100 µm (C, D, F), 1 mm (E), and 20 µm (G–I).

Immunohistochemistry revealed scattered p62-immunoreactive intracytoplasmic inclusions in interneurons with large cell bodies, which could probably be considered Golgi cells, in the cerebellar granular cell layers and in the neurons of the inferior olivary nuclei (Fig. 2G). Very few phosphorylated TAR DNA-binding protein 43 (TDP-43)-immunoreactive threads were observed in the molecular layers of the cerebellum and the uncus of the anterior hippocampus (Fig. 2H). No inclusion immunoreactive signals for expanded polyglutamine stretches were detected.

As a comorbidity, argyrophilic grain disease, which corresponds to Saito stage III disease [16], was observed (Fig. 2I). In particular, argyrophilic grains were prominent in the amygdala, nucleus accumbens, and septal nuclei. The level of Alzheimer’s disease-related neuropathological changes corresponded to a National Institute on Aging-Alzheimer’s Association classification of “low” [17] (Thal phase for amyloidβ plaques: 4 [18], Consortium to Establish a Registry for Alzheimer’s Disease score: B [19], Braak NFT stage: II [20]). No Lewy body-related α-synucleinopathy was identified.

### Characterization of Rab3A R83W

Rab3A is a member of the Rab family of small G proteins. The activity of Rab proteins is cyclically regulated between the GTP-bound active form and the GDP-bound inactive form. GTP-bound Rab proteins interact with specific proteins known as effector proteins, and the Rab–effector protein complex provides each Rab protein with a specific function. Four consensus sequences for GDP/GTP binding and GTPase activities as well as effector domains (i.e., switch I and II regions) are conserved in the Rab family [7]. Given that the R83W mutation is adjacent to the consensus sequence II of Rab3A (Fig. 1D), we examined whether R83W affects the binding of GTP to Rab3A. We performed a GTP-binding assay using a fluorescently labeled GTP analog, Mant-GTP (2’-/3’-O-(N’-methylanthraniloyl) guanosine-5’-O-triphosphate). In this study, we prepared His6-tagged recombinant Rab3A R83W mutant protein and Rab3A wild-type (WT) protein as a control and measured the fluorescence of Mant-GTP bound to the Rab3A R83W mutant or Rab3A WT using a spectrofluorometer. We observed a time-dependent increase in the binding of Mant-GTP to Rab3A R83W, similar to that observed with the Rab3A WT (Fig. 3A). Although the mutant presented a faster binding rate, the binding stoichiometry was the same as that of the wild type. These results indicate that the R83W mutation does not inhibit the binding of GTP to Rab3A; in other words, Rab3A R83W can be activated properly.

**Figure 3.**
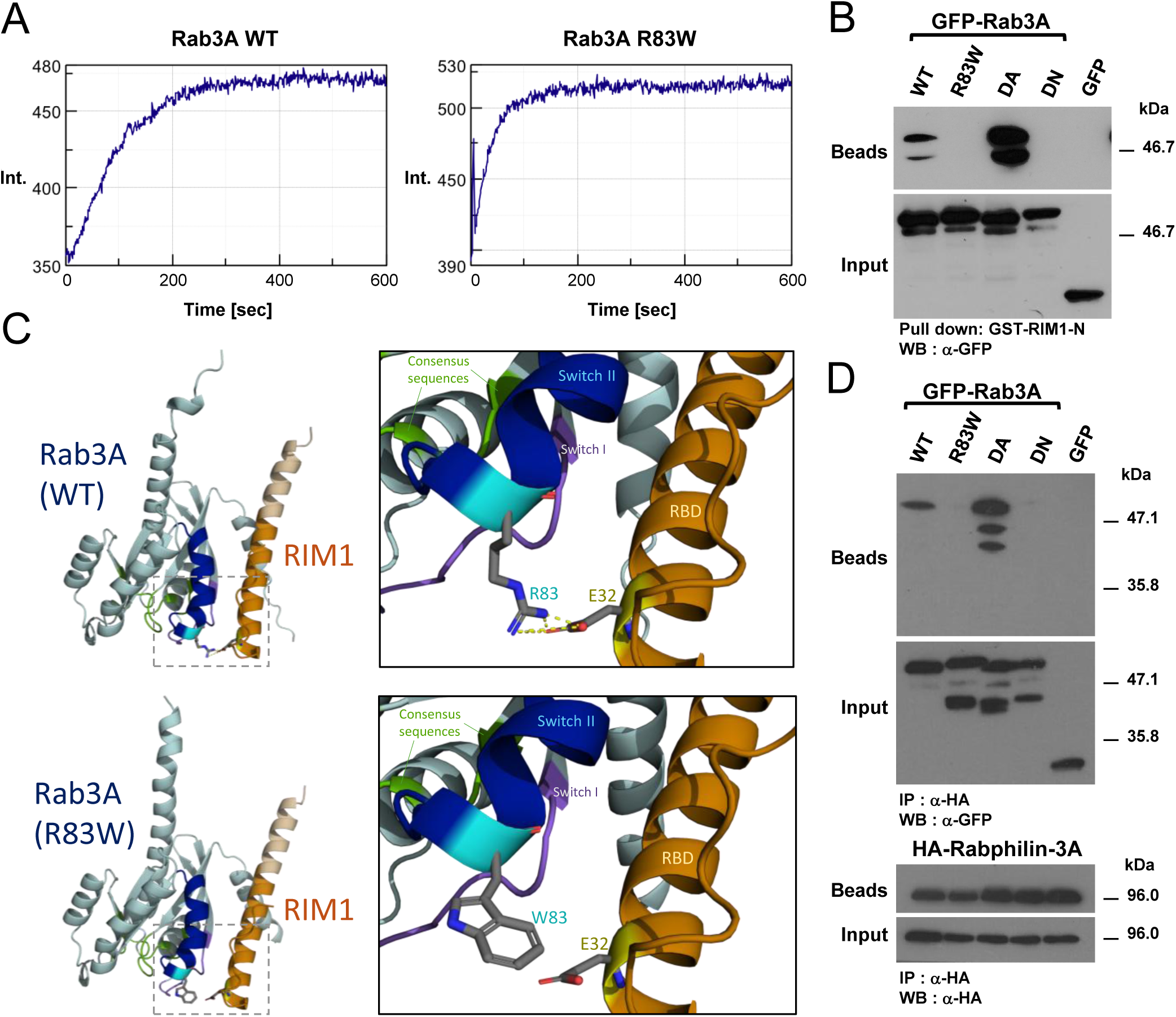
Characterization of Rab3A R83W. (A) *In vitro* exchange assays were performed by measuring the increase in fluorescence intensity (Int.) over time upon incorporation of Mant-GTP into His6-tagged-Rab3A WT or His6-tagged-Rab3A R83W. The data shown are representative of at least three individual experiments. (B) Cell lysates from HEK293 cells expressing GFP-Rab3A WT, GFP-Rab3A R83W, GFP- Rab3A DA, GFP-Rab3A DN, or GFP alone were subjected to pulldown assays using GST-RIM1-N. Pulled-down protein (Beads) and the amount of expressed protein in total cell lysates (Input) were detected by western blotting (WB) with an anti-GFP antibody. (C) Prediction models of the Rab3A–RIM1 complex generated by AlphaFold 3. A salt bridge was observed between the side chain of R83 in Rab3A and the side chain of E32 within the N-terminal short α-helix of RIM1 (indicated by yellow dashed lines). The interaction was disrupted by introducing the R83W mutation. RBD, Rab3A-binding domain. (D) Cell lysates from HEK293 cells expressing GFP-Rab3A WT, mutant (R83W, DA, DN), or GFP alone together with HA-Rabphilin-3A were immunoprecipitated (IP) with an anti-HA antibody. Each immunoprecipitate (Beads) and the amount of expressed protein in total cell lysates (Input) were detected by WB using anti-GFP and anti-HA antibodies.

RIM1 and Rabphilin-3A have been identified as effector proteins for Rab3A [9,10], and the R83W mutation, located within the switch II region, is expected to affect their binding. To examine the effect of R83W on the association of Rab3A with RIM1, we prepared cell lysates expressing GFP-Rab3A WT, GFP-Rab3A R83W, GFP-Rab3A dominant-active (DA) mutant (Q87L), GFP-Rab3A dominant-negative (DN) mutant (T36N), or GFP alone, and then subjected them to a pull-down assay using GST-RIM1- N (aa 1-204), which contains the Rab3A-binding region. GFP-Rab3A DA and GFP- Rab3A WT bound to GST-RIM1-N, but the amount of GFP-Rab3A WT pulled down was less than that of GFP-Rab3A DA (Fig. 3B). However, GFP-Rab3A R83W did not bind to GST-RIM1-N, just as GFP-Rab3A DN and GFP alone did not. Thus, the interaction between Rab3A and RIM1 is likely disrupted by the R83W mutation. We further conducted structural modeling to understand the structural basis of the impaired association between human Rab3A and RIM1. We observed a salt bridge between the side chain of R83 in Rab3A WT and the side chain of E32 in RIM1, and this interaction was predicted to be disrupted by the R83W mutation (Fig. 3C, Fig. S3). We also investigated the effect of R83W on the association of Rab3A with another effector protein, Rabphilin-3A. For this purpose, we prepared HEK293 cells expressing HA-tagged Rab3A WT or the four mutants and then subjected them to an immunoprecipitation assay. HA-Rabphilin-3A was coimmunoprecipitated with GFP- Rab3A DA and GFP-Rab3A WT but not with GFP-Rab3A R83W or GFP-Rab3A DN (Fig. 3D). Notably, the amount of coimmunoprecipitated GFP-Rab3A WT was less than that of GFP-Rab3A DA, which is consistent with the results from the abovementioned pull-down assay. The crystal structure of the Rab3A–Rabphilin-3A complex (PDB: 1ZBD) revealed involvement of the Rab3A residues in Rabphilin-3A binding [21], with R83 being among them (Fig. S4), providing further support for our findings. Taken together, the R83W mutation impairs the formation of both Rab3A–RIM1 and Rab3A– Rabphilin-3A complexes.

### Localization of Rab3A R83W in PC12 cells

Next, to characterize Rab3A R83W not only *in vitro* but also *in cellulo*, we prepared PC12 cells expressing GFP-Rab3A WT, GFP-Rab3A R83W, GFP-Rab3A DA or GFP-Rab3A DN, followed by treatment with nerve growth factor (NGF). In PC12 cells differentiated by NGF, the vesicular staining pattern of GFP-Rab3A WT and GFP- Rab3A DA was observed throughout the cell body, along the processes, and at the tips of the processes as previously reported (Fig. 4) [22, 23]. In contrast, GFP-Rab3A R83W showed a diffuse distribution confined to the cell body, similar to that of GFP-Rab3A DN, which is locked in the GDP-bound form and fails to interact with effector proteins. These results suggest that R83W prevents Rab3A from localizing to neurite tips and being properly targeted to vesicles, likely through the disruption of a Rab3A–effector protein complex.

**Figure 4.**
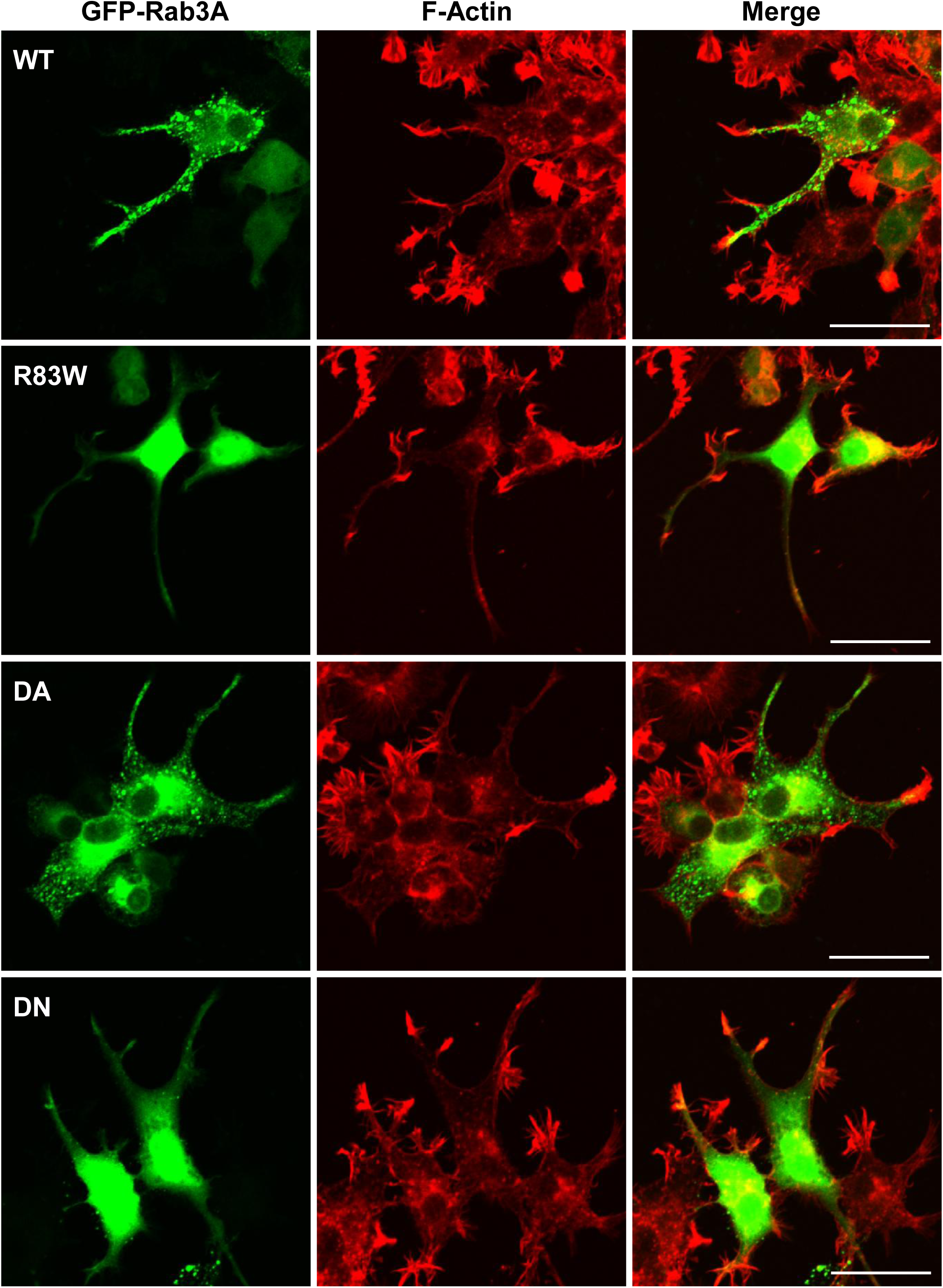
Subcellular localization of Rab3A R83W in PC12 cells. PC12 cells transfected with an expression vector encoding GFP-Rab3A WT, GFP-Rab3A R83W, GFP-Rab3A DA, or GFP-Rab3A DN and treated with NGF were fixed and processed for staining with rhodamine-phalloidin (red). Scale bars: 25 μm.

### Mouse immunohistochemistry

To gain insight into the potential involvement of Rab3A in cerebellar pathology, we investigated the localization of Rab3A in the mouse cerebellum by immunohistochemistry. Rab3A was clearly localized in the molecular layer of the cerebellum, whereas no expression was detected in the dendrites of PCs, which are located in the molecular layer and receive synaptic inputs (Fig. 5A). PFs are long axons of granule cells in the cerebellum, that run horizontally across the molecular layer. They form synapses with the dendrites of PCs, transmitting excitatory signals using glutamate. Rab3A was also colocalized with vesicular glutamate transporter (VGLUT1), a presynaptic marker, indicating its presence at the nerve terminals of PFs (Fig. 5B).

**Figure 5.**
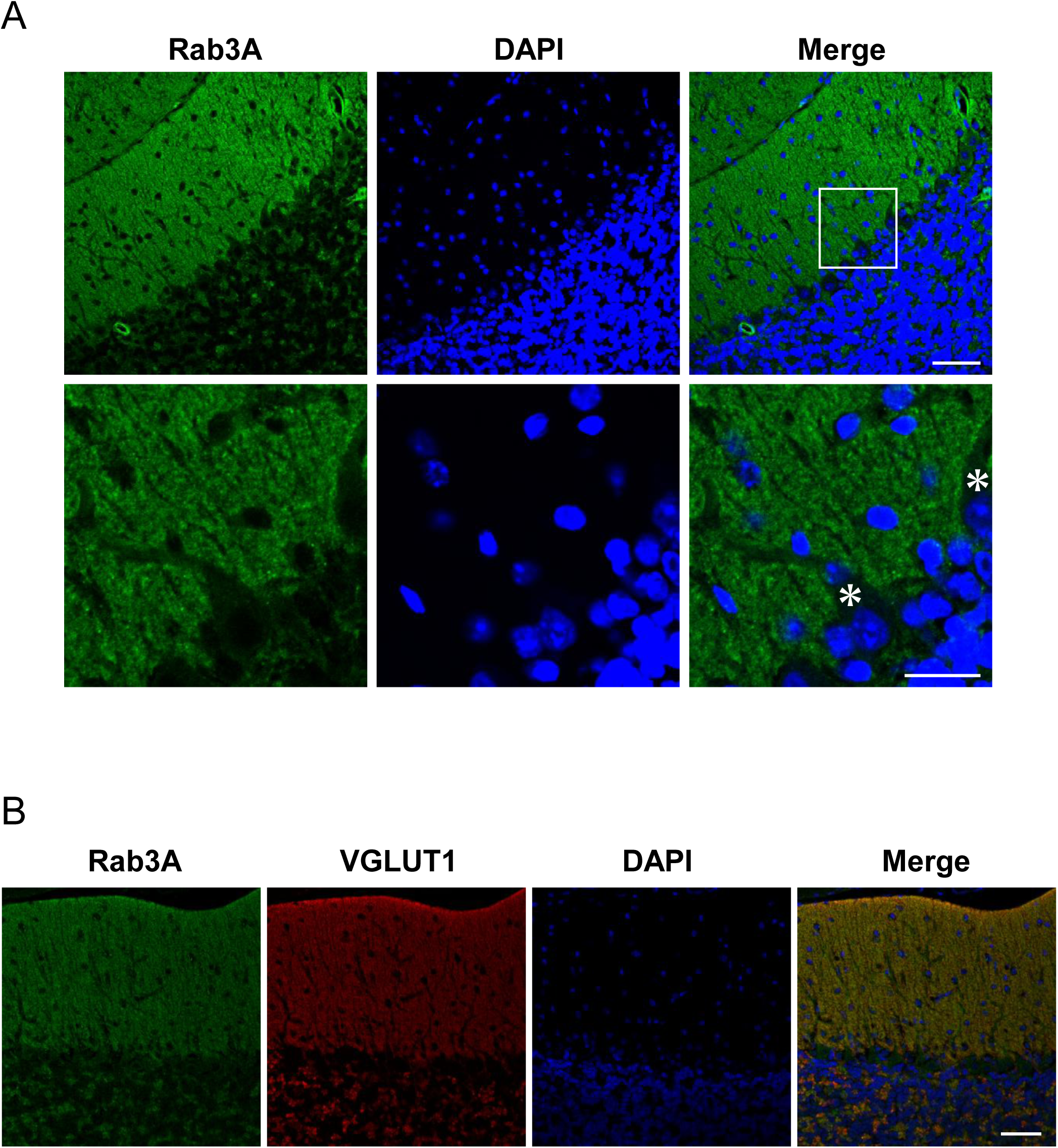
Distribution of Rab3A in the mouse cerebellum. (A) Sections were stained for Rab3A (green) with an anti-Rab3A antibody. The boxed area in the merged image is enlarged (bottom panels). Nuclei were stained with DAPI (blue). Scale bars: 50 µm in the upper panel and 20 µm in the bottom panel. Asterisks indicate Purkinje cells. (B) Sections were double-stained for Rab3A (green) and VGLUT1 (red) using anti-Rab3A and anti-VGLUT1 antibodies. Nuclei were stained with DAPI (blue). Scale bars: 50 µm.

## Discussion

We identified the c.247C>T p.(Arg83Trp) variant in *RAB3A* via whole-exome sequencing in a three-generation pedigree with multiple individuals affected by cerebellar ataxia. The variant showed perfect segregation with the disease, and subsequent genetic screening for the *RAB3A* variant in 320 patients with cerebellar ataxia identified an additional unrelated individual carrying the same variant. During the course of our study, an identical variant in *RAB3A* was independently reported in European families with cerebellar ataxia by Synofzik’s group [24], thereby highlighting the pathogenic significance of the variant and supporting its causative role in SCA.

Clinically, our patients carrying the R83W variant exhibited adult-onset, gradually progressive cerebellar ataxia, frequently accompanied by spasticity, tremors, and neurodevelopmental disorders. The variable combination and severity of these extracerebellar features contributed to intrafamilial phenotypic heterogeneity.

Neuroimaging consistently revealed cerebellar atrophy with predominant vermian involvement. In one patient, pathological examination confirmed diffuse cerebellar cortical degeneration with severe loss of PCs, which was compatible with SCA. These clinical and pathological features suggest that this condition includes both a newly delineated autosomal-dominant ataxia with complex phenotypes, including neurodevelopmental disorders, and vermian-predominant cerebellar atrophy.

The R83 residue, which is altered in affected individuals, is located immediately adjacent to the consensus sequence II involved in GDP/GTP binding and lies within the highly conserved switch II region of Rab3A. The switch regions are particularly important, as they undergo significant conformational changes upon nucleotide binding, forming a critical part of the effector-binding interface [21]. Despite the proximity of the R83 residue to consensus sequence II, the GTP-binding affinity of the R83W mutant protein was preserved in our *in vitro* experiments. Notably, R83W retains intrinsic exchange and hydrolytic activities similar to those of the WT in the recent report by Synofzik’s group [24]. This finding is crucial because it indicates that the R83W mutant is capable of being properly activated and acquiring the GTP-bound conformation, distinguishing it from GTP-binding deficient dominant-negative mutants such as Rab3A T36N [25].

However, critically, we found that the R83W mutation severely impaired the association of Rab3A with its major effector proteins, RIM1 and Rabphilin-3A. Structural modeling provided further insight, revealing a salt bridge between the side chain of R83 in the human Rab3A WT and the side chain of E32 in RIM1. This interaction occurs within the minimal Rab3A-binding domain of RIM1 (aa 19-50), which is sufficient for GTP-dependent binding to Rab3A [26], and the specific contact is predicted to be disrupted by the R83W substitution. Furthermore, on the basis of the crystal structure of the Rab3A–Rabphilin-3A complex, the R83 residue is reported to be among those involved in Rabphilin-3A binding [21]. Therefore, our data, supported by structural information, establish that the R83 residue plays an indispensable role in the protein–protein interactions required for functional engagement with Rab3A effector proteins and that the R83W mutation appears to uncouple the activation state of the protein (GTP-bound) from its ability to interact with these specific effector proteins.

Our experiments demonstrated that the Rab3A WT and the constitutively active GTP-bound mutant formed punctate structures and accumulated at neurite tips, as previously demonstrated [22, 23], mirroring the established role of Rab3A in regulating the dynamics and exocytosis of both SVs and dense-core vesicles. In contrast, the Rab3A R83W mutant exhibited diffuse cytoplasmic localization, similar to that observed with the dominant-negative Rab3A mutant locked in the GDP-bound form.

The concordant localization pattern suggests that R83W-induced mislocalization results from a failure to engage downstream effector proteins, despite preserved GTP binding. In other words, effector protein interaction—enabled by the GTP-bound state—is a key determinant for proper Rab3A targeting, and this process is disrupted by the R83W mutation.

A primary mechanism mediating the localization and function of Rab3A, particularly its tethering role at the active zone, involves its interaction with the active zone scaffold protein RIM1 [27, 28]. Rab3A localization is known to be independent of Rabphilin-3A based on observations from Rabphilin-3A-deficient mice [29]. Moreover, expression of the Rab3A point mutant T54A, which is unable to bind Rabphilin-3A, in PC12 cells revealed that the mutant protein localizes to secretory vesicles similar to the wild-type protein [23]. These findings collectively suggest that the mislocalization of Rab3A R83W may be more attributable to disruption of the Rab3A–RIM1 complex than to the inability to bind Rabphilin-3A.

In this study, we confirmed that Rab3A expression in the cerebellum is highly restricted to the molecular layer, which is consistent with previous reports [29, 30]. While these earlier studies did not describe Rab3A expression in PCs in detail, our experiments demonstrated that Rab3A is scarcely expressed in PCs, which are the postsynaptic targets of PFs. This finding aligns with the observed colocalization of Rab3A with VGLUT1, a specific presynaptic marker enriched at PF terminals [31]. PFs form numerous synaptic contacts on PCs, enabling fine-tuned modulation of PC activity [32]—a process involving the Rab3A–RIM1 complex [33]. Alterations of the PF–PC synapse have been implicated in several experimental models of SCA [5, 32], and specifically, the knockdown of FGF14 in granule cells leads to reduced Ca^2+^ currents and impaired vesicular recycling [34]. Moreover, a recent study on AP-2, which is essential for clathrin-mediated SV endocytosis, revealed that an imbalance between PF and climbing fiber signals resulting from impaired vesicle recycling leads to hyperexcitability and degeneration of PCs [35]. These findings highlight that PF–PC synapses, with their sustained high-frequency activity and heavy vesicle recycling load [36], are especially susceptible to stress from synaptic protein defects. Similarly, the Rab3A R83W mutation might induce activity-dependent stress, compromise PF terminal function, and ultimately trigger neurodegeneration.

Features of neurodevelopmental disorders, ranging from ASD traits to intellectual disability, were frequently observed in affected individuals with the R83W variant. Accumulating evidence suggests that the cerebellum is involved in cognition, learning and affective regulation in addition to its role in motor control, and cerebellar dysfunction is clearly evident in several developmental disorders, including autism [37]. While neurodevelopmental disorders are frequently associated with autosomal-recessive cerebellar ataxias [38], several autosomal-dominant forms caused by *de novo* or dominantly inherited variants have also been linked to ASD, intellectual disabilities, or global developmental delay [39, 40]. The constellation of motor symptoms and features of neurodevelopmental disorders observed in our patients further indicates that the pathogenic impact of the R83W mutation extends beyond the cerebellar motor circuit.

In support of this interpretation, recent studies have increasingly emphasized the importance of synaptic dysfunction in the etiology of neurodevelopmental disorders [41]. Importantly, Rab3A and RIM1 function as general modules for long-term synaptic plasticity, which is implicated in diverse forms of learning and memory [42] and operates at cerebellar PF and hippocampal mossy fiber synapses [43]. Moreover, both Rab3A-deficient and RIM1 knockout mice exhibit impairments in learning and memory tasks [44, 45]. These findings underscore that the Rab3A–RIM1 axis is a key mediator of synaptic plasticity in cognitive domains, and suggest that its disruption by the R83W variant may underlie neurodevelopmental abnormalities.

In summary, we identified the R83W variant in Rab3A in two families that presented with adult-onset cerebellar ataxia accompanied by various features of neurodevelopmental disorders. A functional study suggested that disruption of the Rab3A–RIM1 complex at PF–PC synapses might represent a contributing pathological mechanism underlying this newly identified form of SCA. Our findings provide a foundation for future therapeutic strategies targeting Rab3A or its associated pathways, especially in light of growing evidence that synaptic dysfunction is a common pathological mechanism in various SCAs and neurodevelopmental disorders.

## Materials and Methods

### Clinical assessment

Clinical assessments were carried out at Tokushima University Hospital, Itsuki Hospital, and National Center Hospital by board-certified neurologists (RM, TF, RO, YO, YN, YT, and YI). All affected individuals except for T-I:2 underwent routine clinical evaluations, including history acquisition, neurological examination, blood testing, cognitive assessment using the Mini-Mental State Examination (MMSE), and brain MRI. Clinical information for T-I:2 was obtained through an interview with a healthy family member (T-II:4). Information on neurodevelopmental disorders and related features was obtained from medical records and supplemented by patient or caregiver reports. Written informed consent was obtained prior to video-recording of the neurological symptoms of the affected individuals.

### Genome-wide linkage analysis

Genomic DNA (gDNA) was extracted from the peripheral lymphocytes of the participants. All procedures were approved by the Ethical Review Boards of Tokushima University and Hiroshima University. Written informed consent was obtained from all participants prior to enrollment. After confirming that the proband (T-II:2) tested negative for pathogenic repeat expansions associated with SCA1, SCA 2, SCA 3, SCA 6, SCA 7, SCA 15, SCA 17, SCA 31, and SCA 36, six individuals from Family T (II:1, II:2, II:4, II:5, II:7, and III:3) were genotyped using the Infinium Asian Screening Array-24 v1.0 BeadChip Kit, which contains 642,824 markers (Illumina, San Diego, CA, USA).

Multipoint parametric linkage analysis was carried out with Merlin-1.1.2 with a dominant inheritance model assuming complete penetrance and a disease allele frequency of 0.0001 at three equally spaced locations between each consecutive pair of markers.

### Whole-exome analysis

Whole-exome sequencing was performed on 6 individuals in Family T (II:1, II:2, II:4, II:5, II:7 and III:3). gDNA libraries were prepared using SeqCap EZ Human Exome Library v3.0 (Roche NimbleGen, Madison, WI). Sequencing was conducted using 100-bp paired-end reads on a HiSeq1500 sequencer (Illumina). The reads were aligned to the human reference genome using the Burrows–Wheeler Aligner (BWA) [46], and variants were called using the Genome Analysis Toolkit (GATK) [47]. Variant annotation was performed with ANNOVAR [48] using the dbSNP Build 135, 1000 Genomes Project, and ESP5400 databases. Sequencing depth was calculated using GATK. In-house control exomes were obtained from 135 individuals who underwent whole-exome analysis. The pathogenicity of variants was assessed using Combined Annotation Dependent Depletion (CADD) [49], Sorting Intolerant From Tolerant (SIFT) [50], PolyPhen-2 [51], and MutationTaster [52]. The identified variant was validated in both affected and unaffected individuals via PCR-based amplification followed by Sanger sequencing using the Applied Biosystems 3130 Genetic Analyzer (Applied Biosystems, Foster City, CA, USA).

### Amplicon sequencing

We screened individuals with SCA for *RAB3A* variants, including 225 individuals with autosomal-dominant inheritance, 55 with autosomal-recessive inheritance, and 40 with sporadic cases, using the Ion Proton System (Life Technologies, Carlsbad, CA, USA). Primers targeting the coding region of *RAB3A* were designed using the AmpliSeq Designer (https://www.ampliseq.com/). Library preparation was carried out with the Ion AmpliSeq Library Kit 2.0 (Life Technologies) following the manufacturer’s protocol. Template preparation and chip loading were performed using the Ion Chef system, and sequencing was conducted on an Ion Proton sequencer with an Ion PI chip (Life Technologies). Variant calling was performed using Torrent Suite software 5.10.

### Neuropathological investigation

A brain autopsy was performed for patient T-II:1. The left half of the brain was fixed in 20% buffered formalin, while the right half was sliced and frozen. The right cerebral hemisphere, brainstem, and cerebellum were then dissected on the coronal, axial, and sagittal planes, respectively. Representative anatomical areas were embedded in paraffin, and 6-µm-thick sections were subjected to hematoxylin and eosin staining, Klüver-Barrera staining, and Gallyas–Braak silver impregnation. Subsequently, immunoreaction product deposits on immunohistochemically stained sections were visualized with a Ventana BenchMark GX autostainer (Ventana Medical Systems, Tucson, AZ, USA), an I-View Universal DAB Detection Kit (Roche Diagnostics, Basel, Switzerland), and primary antibodies against p62 (mouse monoclonal; clone 3/P62 LCK LIGAND; BD Biosciences, San Jose, CA, USA), amyloid β (mouse monoclonal; clone 12B2; IBL, Gunma, Japan), phosphorylated TDP43 (mouse monoclonal; clone 11-9, Cosmo Bio, Tokyo, Japan), phosphorylated neurofilament (mouse monoclonal; clone SMI31; Sternberger Monoclonals Inc., Baltimore, MA, USA), expanded polyglutamine stretches (mouse monoclonal; clone 5TF1-1C2; Millipore, Billerica, MA, USA), phosphorylated tau (mouse monoclonal; clone AT8; Innogenetics, Ghent, Belgium), and phosphorylated α-synuclein (mouse monoclonal; clone pSyn#64, FUJIFILM Wako Pure Chemical Corporation, Osaka, Japan). The samples were examined with a light microscope (Eclipse Ni, Nikon, Tokyo, Japan) and photographed using a digital camera (DS-Ri2, Nikon).

### Structural modeling and visualization of Rab3A complexes

The predicted structures of human Rab3A (UniProtKB: P20336) and the Rab3A-binding domain of human RIM1 (aa 19–50; UniProtKB: Q86UR5) were generated using AlphaFold 3 (v3.0.1, https://alphafoldserver.com/) [53] to investigate the structures of the Rab3A–RIM1 complexes. For both the wild-type and the R83W mutant forms of Rab3A, five models were generated and ranked by AlphaFold’s internal confidence metric (ranking_score). The highest-confidence model for each complex was selected for further analysis. Structural alignment was performed by superimposing the five predicted models with Rab3A as the alignment reference, allowing assessment of structural consistency across predictions. In addition, the crystal structure of the Rab3A–Rabphilin-3A complex (PDB ID: 1ZBD) [21] was used to show the structural context of the R83 residue. Both the predicted and crystal structures were visualized using PyMOL (v2.5.0 Open-Source, https://github.com/schrodinger/pymol-open-source). To assess interactions between Rab3A residue 83 and RIM1 residue E32, interatomic distances were measured using a 4.0 Å cutoff.

### Plasmid construction

pEGFP-bovine Rab3A R83W was constructed using a QuikChange Lightning Multi Site-Directed Mutagenesis Kit (Agilent Technologies Inc., Santa Clara, CA, USA). The cDNAs encoding Rab3A WT, Rab3A Q87L (constitutively active form), Rab3A T36N (constitutively negative form), and Rab3A R83W were subcloned and inserted into pEGFP or pCMVHA. To generate His6-tagged proteins, the cDNAs of Rab3A WT and Rab3A R83W were subcloned and inserted into pRSET-A. RIM1-N (aa 1-204) cDNA was amplified by PCR from pCMVTag3B-hRIM1, which was a kind gift from Dr. Susumu Seino, and subcloned and inserted into pGEX6P-1. The cDNA encoding Rabphilin-3A was subsequently subcloned and inserted into pCIneo-Myc. All the plasmids constructed in this study were sequenced using an ABI Prism 3100 genetic analyzer (Applied Biosystems).

### Antibodies

The mouse monoclonal anti-Rab3A antibody (clone SG-11-7) was prepared as previously described [54]. Rabbit anti-VGLUT1 was purchased from Merck Millipore (Burlington, MA, USA). The mouse anti-Myc antibody (clone 9E10) was produced from a hybridoma obtained from ATCC (Manassas, VA, USA); the mouse anti-hemagglutinin (HA) (clone 12CA5) and rat anti-HA (clone 3F10) antibodies were from Roche Diagnostics; the mouse anti-GAPDH antibody was from Applied Biosystems; the rabbit anti-GFP antibody, and rhodamine-phalloidin were from Invitrogen (Carlsbad, CA, USA); and the Alexa 488- or Cy3-conjugated secondary antibodies and HRP-coupled secondary antibodies were obtained from Invitrogen and Jackson ImmunoResearch Laboratories (West Grove, PA, USA), respectively.

### Cell culture and transfection

PC12 cells were cultured in Dulbecco’s modified Eagle’s medium (DMEM) supplemented with 10% fetal bovine serum (FBS), 5% horse serum, 100 U/ml penicillin, and 100 μg/ml streptomycin and were maintained at 37°C in a water-saturated atmosphere of 90% air and 10% CO_2_. HEK293 cells were cultured in DMEM with 5% FBS, 1 mM sodium pyruvate, 100 U/ml penicillin, and 100 μg/ml streptomycin and were maintained at 37°C in a water-saturated atmosphere of 95% air and 5% CO_2_. PC12 cells and HEK293 cells were transfected with Lipofectamine 2000 transfection reagent (Invitrogen) and PEI-MAX transfection reagent, respectively.

### Bacterial production of recombinant proteins

The GST-fusion protein (GST-RIM1-N) was expressed in the *Escherichia coli* (*E. coli*) strain DH5α cultured in the presence of 0.1 mM isopropyl-β-D-thiogalactopyranoside (IPTG). Bacteria were resuspended in Buffer B [20 mM Tris-HCl (pH 7.5), 1 mM EDTA, 1 mM EGTA, and 1 mM dithiothreitol] containing 10 μM p-amidinophenyl methanesulfonyl fluoride (APMSF), and 1 μg/ml leupeptin and lysed by sonication; the resulting cellular debris was removed by centrifugation for 60 min at 100,000 × *g*. The resulting supernatant was applied to a Glutathione-Sepharose 4B column (Cytiva, Marlborough, MA, USA). The pass-through fraction was applied a second time to the same column. The column was washed with Buffer B, and the GST fusion proteins were eluted in Buffer C [50 mM Tris-HCl (pH 8.0), 10 mM reduced glutathione] and dialyzed with Buffer B. The purified protein was detected by electrophoresis and CBB staining (data not shown). The His6-tagged proteins were expressed in the *E. coli* strain BL21 (DE3) pLysS (Novagen, EMD Biosciences, Inc., San Diego, CA, USA), which was subsequently cultured in the presence of 0.1 mM IPTG. Bacteria were resuspended in Buffer C [20 mM Tris-HCl (pH 8.0), 100 mM NaCl, and 5 mM MgCl_2_] containing 10 μM APMSF, and lysed by sonication. Recombinant proteins were purified using TALON Metal Affinity Resin (Clontech Laboratories, San Jose, CA, USA) according to the manufacturer’s recommendations with slight modifications.

### Pull-down assay

HEK293 cells were seeded in 60-mm dishes and transfected the following day with an appropriate amount of each plasmid using the PEI-MAX transfection reagent. After 48 h of incubation, the cells were lysed and then centrifuged to remove debris.

Each supernatant was incubated with purified GST-RIM1-N attached to glutathione– Sepharose beads. The beads were then washed and resuspended in sodium dodecyl sulfate (SDS) sample buffer. Comparable amounts of the proteins that remained associated with the beads were separated by SDS–polyacrylamide gel electrophoresis (PAGE). The fraction of target protein bound to the beads was determined by western blotting using an anti-GFP antibody (Invitrogen) as described.

### Immunoprecipitation

Forty-eight hours after transfection, HEK293 cells were lysed and then centrifuged to remove debris. An aliquot of the supernatant was used to verify the expression of the indicated proteins. The remaining supernatant was mixed with Protein G–Sepharose FF beads (GE Healthcare Biosciences, Chicago, IL, USA) linked to an anti-HA monoclonal antibody (12CA5; Roche Diagnostics). The beads were washed and then resuspended in SDS sample buffer. The immunoprecipitates were subjected to western blotting with anti-HA (3F10; Roche Diagnostics) and anti-Myc (9E10) antibodies as previously described.

### Immunoblotting

The cell lysates, immunoprecipitates, or pulled-down proteins were separated by SDS[PAGE, transferred to Immobilon-P membranes (Merck Millipore), and blocked for 1 h in Tris-buffered saline containing 5% nonfat dry milk. After incubation with specific primary antibodies for 1 h, followed by incubation with HRP-coupled secondary antibodies (Jackson) for 1 h, immunoreactive proteins were visualized using Western Blotting Substrate Plus (Pierce, Thermo Fisher Scientific, Waltham, MA, USA).

### Guanine nucleotide binding assay

2’Deoxy-3’O-N-methylanthraniloyl GTP (Mant-GTP) is a fluorescent nucleotide GTP analog (Abcam). Rab3A WT and Rab3A R83W (3.6 μM final concentration) were mixed with Mant-GTP (10 μM final concentration) in reaction buffer (20 mM Tris-HCl at pH 7.5, 50 mM NaCl, 1 mM DTT, 0.5 mM MgCl_2_, and 5 mM EDTA). The mixtures were then directly subjected to spectral measurement on an FP-6300 spectrofluorometer (Jasco, Tokyo, Japan) with excitation at 433 nm and 30°C for 10 min. The Mant-GTP binding was plotted in arbitrary units for fluorescence over time (excitation at 361 nm, emission at 446 nm).

### Immunocytochemistry

PC12 cells were seeded at a density of 4 × 10^5^ cells on poly-L-lysine-coated coverslips in 35-mm dishes and transfected the following day with various DNA plasmids. After 48 h of incubation, the cells were fixed with 4% formaldehyde in PBS at room temperature for 20 min. The cells were then incubated with 5% donkey serum and 0.1% Triton X-100 in PBS for 30 min. F-actin was labeled with rhodamine phalloidin (Invitrogen). The stained cells were observed under an A1 confocal laser-scanning microscope (Nikon).

### Mouse immunohistochemistry

Mice were anesthetized with ether and subjected to vascular perfusion with 4% paraformaldehyde in 0.1 M phosphate buffer, pH 7.4 (PB). The cerebellum was cut into slices and further fixed by immersion in the same fixative at 4°C overnight. The fixed cerebellum was cryoprotected by sequentially immersing it in 12%, 15%, and 18% sucrose in 0.1 M PB, embedding it in O.C.T. compound (Sakura Fine Technical Co. Ltd.), and then quickly freezing it in liquid nitrogen. Five-micrometer-thick frozen sections were prepared with a Leica CM 1850 cryostat and mounted on silane-coated glass slides. The sections were incubated with 17% goat serum in dilution buffer (450 mM NaCl and 0.3% Triton X-100 in 20 mM PB) for 30 min, followed by incubation with primary antibodies for 2 h. After washing with dilution buffer, the sections were incubated with secondary antibodies for 1 h. The nuclei were labeled with 4,6- diamidine-2-phenylindole dihydrochloride (DAPI) (KPL Inc., Gaithersburg, MD, USA). The sections were mounted in Aqua-Poly/Mount (Polysciences, Inc., Warrington, PA, USA), and the samples were observed using an A1 confocal laser scanning microscope (Nikon).

## Acknowledgement

This work was supported by MHLW Research on ataxia (Program Grant Number JPMH23FC1010) and the Rare and Intractable Diseases Program (Grant Number JPMH23FC1008). We thank Yoshimi Mihara (Department of Neurology, Tokushima University Graduate School of Biomedical Sciences) for her technical assistance.

## Data Availability

The data are available upon reasonable request from the corresponding author.

