## Supplementary information for "An R83W mutation in Rab3A causes autosomal-dominant cerebellar ataxia"

1. **Supplementary Figure S1.** Brain MR images of the affected individuals

2. **Supplementary Figure S2.** Results of linkage analysis

3. **Supplementary Table S1.** Sequence metrics and variant filtering

4. **Supplementary Figure S3.** Structural analysis of the human Rab3A–RIM1 complex by AlphaFold 3

5. **Supplementary Figure S4.** Structural interface of the Rab3A–Rabphilin-3A complex (PDB: 1ZBD) involving residue R83

1. **Supplementary Figure S1.** Brain MR images of the affected individuals

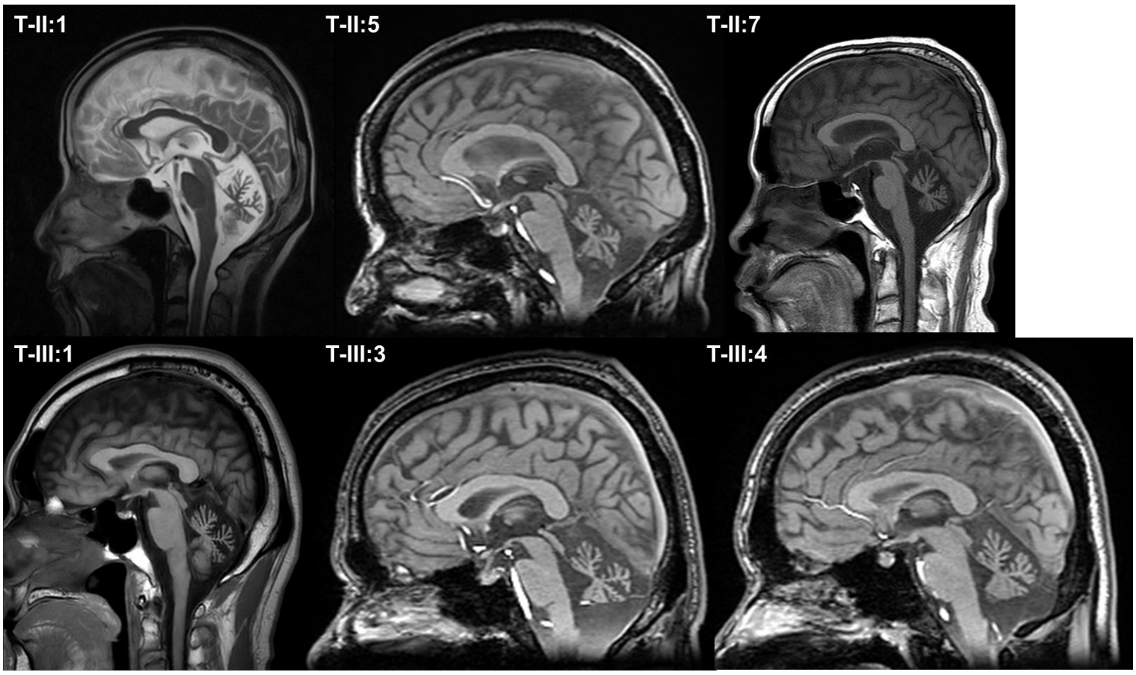


T-II:1, sagittal T2-weighted image; T-II:5, T-III:4, and T-III:6, sagittal T1-weighted 3D gradient echo images; T-II:7 and T-III:1, sagittal T1-weighted images. Cerebellar vermian atrophy in affected individuals in Family T.

2. **Supplementary Figure S2.** Results of linkage analysis

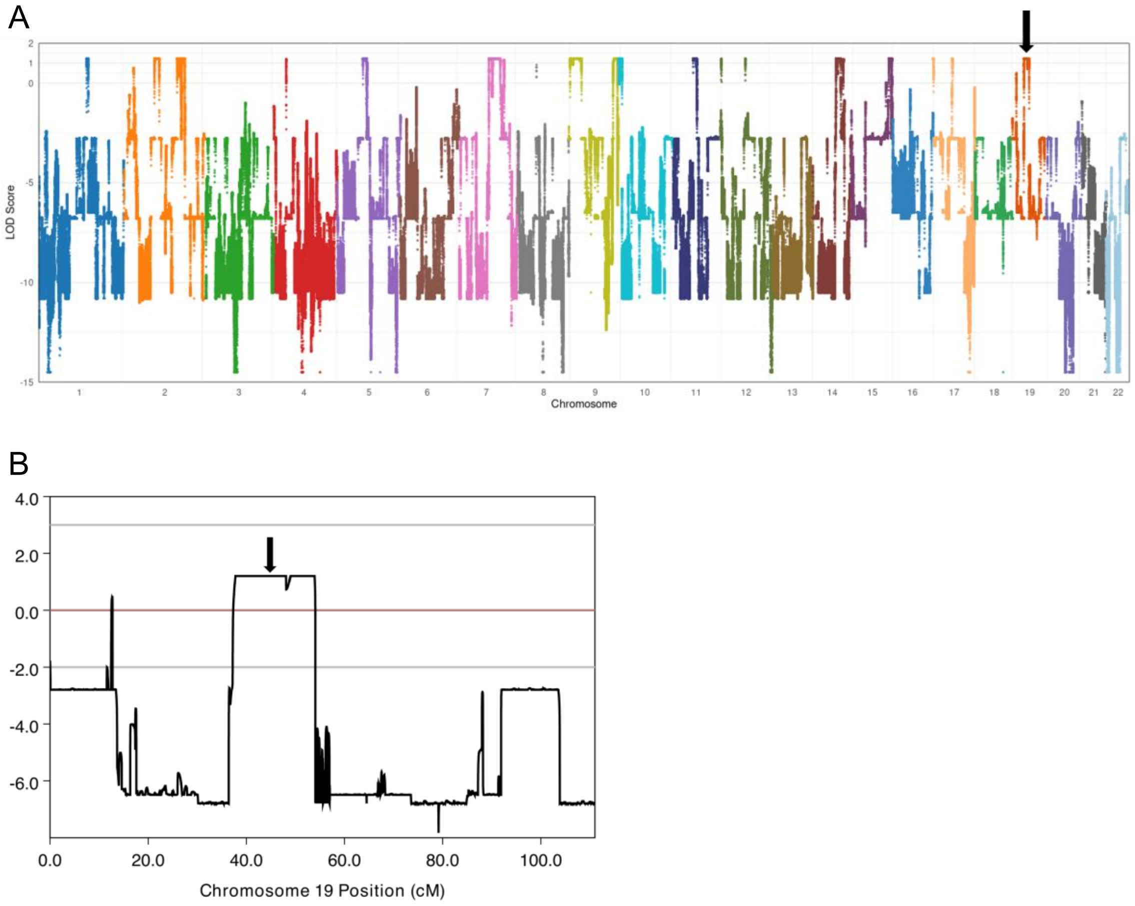


(A) Multipoint LOD scores across chromosomes 1–22. (B) Enlarged view of chromosome 19 showing the detailed distribution of LOD scores along its genetic map. The arrows in (A) and (B) indicate the position of *RAB3A*. One of the regions with the highest LOD score was located on chromosome 7 and overlapped with the previously reported locus for SCA18 (MIM: 607458). However, no compelling variants were identified in this region by exome sequencing, and the clinical phenotype of the affected individuals was distinct from that of SCA18, which is characterized by sensorimotor neuropathy.

3. **Supplementary Table S1.** Sequence metrics and variant filtering

| | T-II:1 | T-II:2 (proband) | T-II:4 (unaffected) | T-II:5 | T-II:7 | T-III:4 |
|---|---|---|---|---|---|---|
| Total reads | 81,343,646 | 80,040,536 | 83,022,748 | 158,647,190 | 90,503,828 | 160,469,498 |
| Mappable reads | 77,246,095 | 76,171,008 | 78,673,051 | 145,386,823 | 85,917,575 | 147,161,610 |
| % Mappable reads (out of total reads) | 94.9 | 95.2 | 94.8 | 91.6 | 94.9 | 91.7 |
| Mean read depth of target regions | 59× | 58× | 61× | 116× | 66× | 117× |
| Filters applied | Number of variants | | | | | |
| Total variants | 63, 074 | 63,158 | 63,056 | 64,928 | 63,201 | 64,769 |
| Heterozygous variants | 39,022 | 39,403 | 39,105 | 40,849 | 38,838 | 40,773 |
| Nonsynonymous/Frameshift/Stop/Splicing variants | 5,497 | 5,561 | 5,649 | 5,574 | 5,562 | 5,585 |
| Not present in dbSNP135, 1000 genomes, and ESP5400 | 198 | 189 | 222 | 218 | 218 | 210 |
| Not present in in-house database | 105 | 87 | 113 | 105 | 113 | 110 |
| CADD phred > 20 | 56 | 44 | 59 | 58 | 61 | 52 |
| Segregate with ataxia phenotype | 1 (NM_002866.5 (***RAB3A***):c.247C>T p.(Arg83Trp)) | | | | | |

Whole-exome sequencing metrics and results of stepwise variant filtering for six individuals from Family T, including five affected individuals and one unaffected individual. Variant prioritization led to the identification of a single heterozygous *RAB3A* variant that fully segregated with the ataxia phenotype.

Abbreviations: CADD, combined annotation-dependent depletion; ESP5400, Exome Sequencing Project (5400 exomes); dbSNP, single-nucleotide polymorphism database.

4. **Supplementary Figure S3.** Structural analysis of the human Rab3A–RIM1 complex by AlphaFold 3

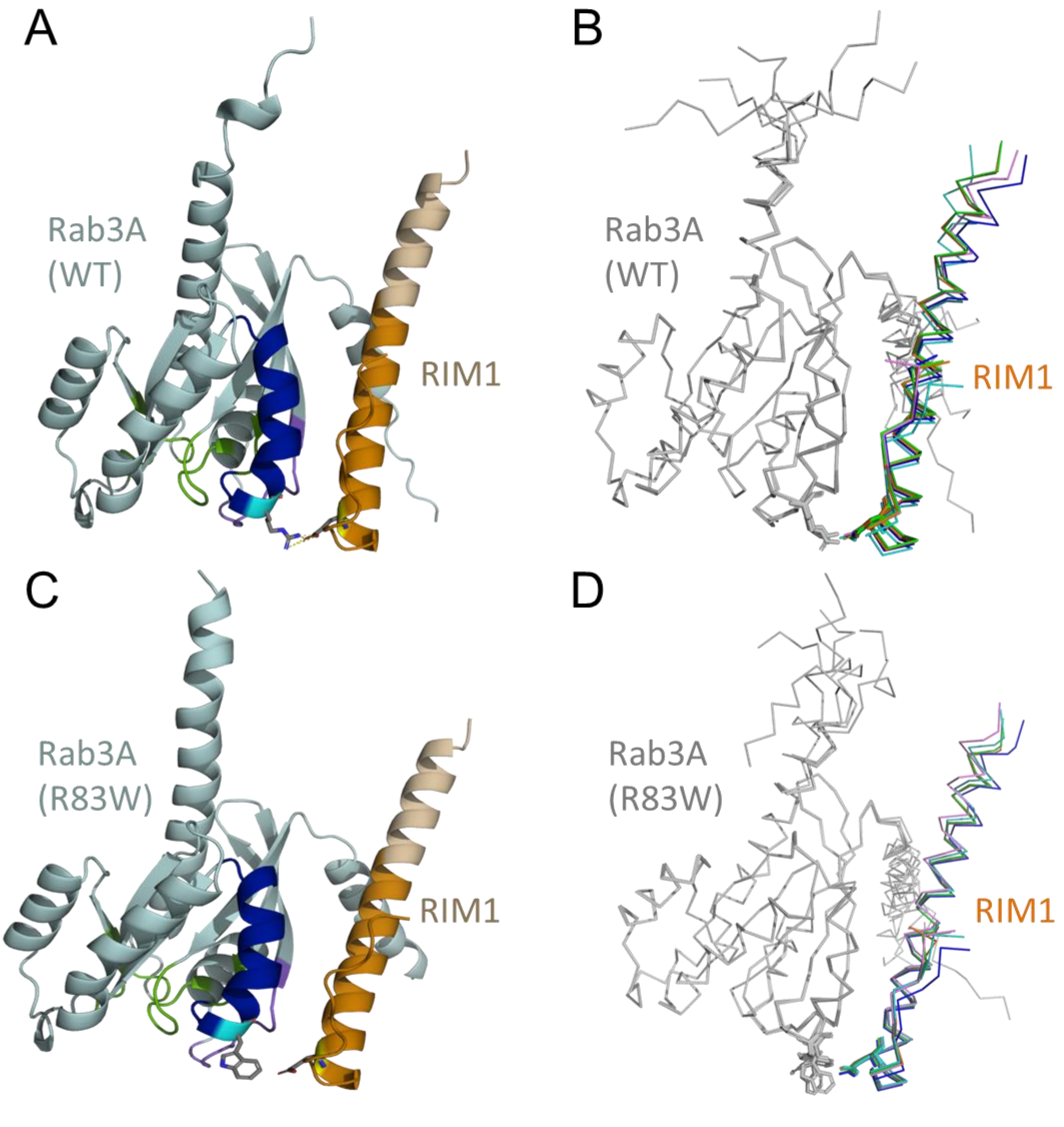


Structural modeling of the Rab3A WT–RIM1 (A, B) and Rab3A R83W–RIM1 (C, D) complexes generated using AlphaFold3. Panels A and C show the top-ranked prediction model for each complex. Panels B and D display all five top-ranked models aligned based on the structure of Rab3A. The models include Rab3A residues 1–203 and RIM1 residues 19–50, which correspond to the Rab3A-binding region. In both B and D, the predicted structures of Rab3A and RIM1, as well as their relative positions, were largely consistent across the models. These findings suggest that the core interaction between Rab3A and RIM1 was modeled in a stable and reproducible manner.

Abbreviations: WT, wild-type.

5. **Supplementary Figure S4.** Structural interface of the Rab3A–Rabphilin-3A complex (PDB ID: 1ZBD) involving residue R83

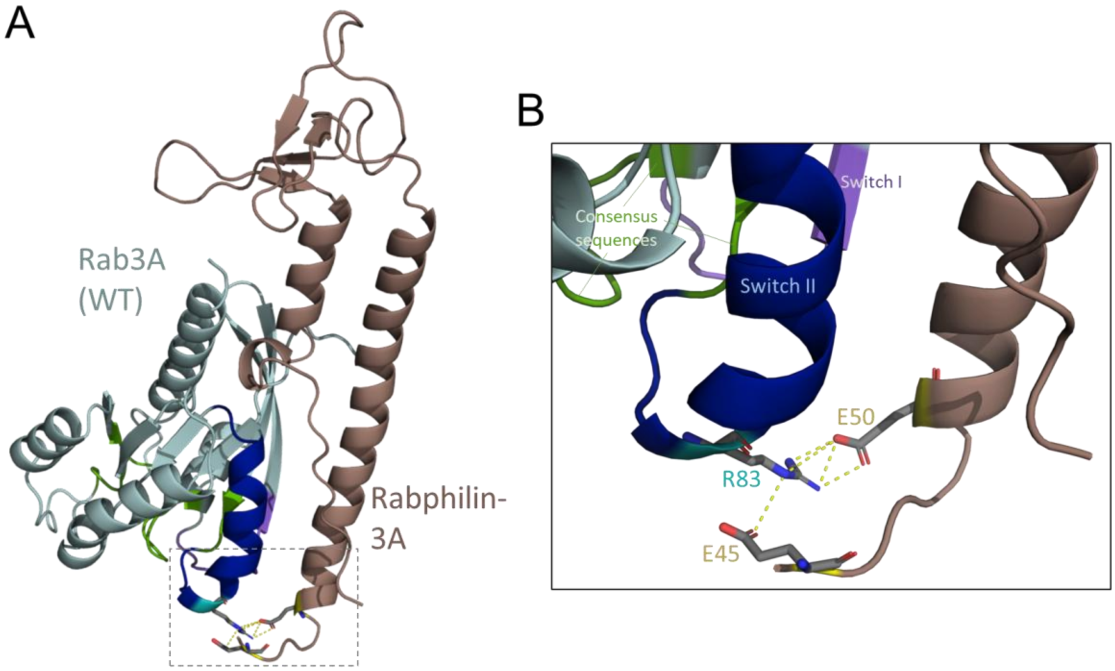


(A) Overall structure of the Rab3A–Rabphilin-3A complex. (B) Close-up view focusing on R83 of Rab3A and its proximity to E45 and E50 of Rabphilin-3A. R83 lies within 4.0 Å of E45 and E50 of Rabphilin-3A, as indicated by yellow dashed lines representing interatomic distances. These contacts illustrate the involvement of R83 in the protein–protein interface, as previously described [1].
